# Moderate-Intensity Exercise Versus High-Intensity Interval Training to Recover Walking Post-Stroke: Protocol for a Randomized Controlled Trial

**DOI:** 10.1101/2021.06.25.21259562

**Authors:** Allison Miller, Darcy S. Reisman, Sandra A. Billinger, Kari Dunning, Sarah Doren, Jaimie Ward, Henry Wright, Erin Wagner, Daniel Carl, Myron Gerson, Oluwole Awosika, Jane Khoury, Brett Kissela, Pierce Boyne

## Abstract

**Background:** Stroke results in neurologic impairments and aerobic deconditioning that contribute to limited walking capacity which is a major barrier post-stroke. Current exercise recommendations and stroke rehabilitation guidelines recommend moderate-intensity aerobic training post-stroke. Locomotor high-intensity interval training is a promising new strategy that has shown significantly greater improvements in aerobic fitness and motor performance than moderate-intensity aerobic training in other populations. However, the relative benefits and risks of high-intensity interval training and moderate-intensity aerobic training remain poorly understood following stroke. In this study, we hypothesize that locomotor high-intensity interval training will result in greater improvements in walking capacity than moderate-intensity aerobic training.

**Methods:** Using a single-blind, 3-site randomized controlled trial, 50 chronic (>6 months) stroke survivors are randomly assigned to complete 36 locomotor training sessions of either high-intensity interval training or moderate-intensity aerobic training. Main eligibility criteria are: age 40-80 years, single stroke for which the participant received treatment (experienced 6 months to 5 years prior to consent), walking speed ≤1.0 m/s, able to walk at least 3 minutes on the treadmill at ≥ 0.13 m/s (0.3 mph), stable cardiovascular condition (American Heart Association class B), and the ability to walk 10 meters overground without continuous physical assistance. The primary outcome (walking capacity) and secondary outcomes (self-selected and fast gait speed, aerobic fitness and fatigue) are assessed prior to initiating training and after 4 weeks, 8 weeks, and 12 weeks of training.

**Discussion:** This study will provide fundamental new knowledge to inform the selection of intensity and duration dosing parameters for gait recovery and optimization of aerobic training interventions in chronic stroke. Data needed to justify and design a subsequent definitive trial will also be obtained. Thus, the results of this study will inform future stroke rehabilitation guidelines on how to optimally improve walking capacity following stroke.

**Trial Registration:** ClinicalTrials.gov Identifier: NCT03760016. First posted: November 30, 2018. https://clinicaltrials.gov/ct2/show/NCT03760016

## BACKGROUND

### Background and Rationale

Approximately 6.6 million Americans are currently living with chronic sequelae of stroke of which a primary impairment is reduced walking capacity (1). Limited walking capacity is a major barrier to recovery after stroke (2), and less than 10% of stroke survivors have adequate walking speed and endurance to allow for normal daily functioning, such as grocery shopping and occupational requirements (1, 3-6). Thus, improving walking capacity is a primary goal of rehabilitation after stroke (2, 7).

To address impairments in walking capacity, current exercise recommendations and stroke rehabilitation guidelines recommend moderate-intensity aerobic training (MAT) (2, 8). Compared to conventional rehabilitation approaches and lower intensity training, MAT has shown significant benefits across a range of outcomes, such as improvements in aerobic fitness (9-11), walking capacity (10-14), and overall disability (14). However, this approach has known limitations that has restricted its adoption in most clinical stroke rehabilitation settings (2). In particular, MAT has shown small and inconsistent effects on gait speed, a primary outcome of stroke rehabilitation (10, 11, 13). In addition, most laboratory-based MAT protocols have involved training durations (typically 45 minutes, 3x/week for 6 months) (15-22) beyond what is possible in clinical practice due to issues related to patient adherence (23-25) and reimbursement (26, 27). Thus, to improve walking capacity post stroke, there is a critical need for a more efficacious and time-efficient intervention.

Recent evidence suggests that a more vigorous training intensity (>60% vs. 40-60% heart rate reserve) may be a ‘critical ingredient’ for greater and more rapid improvements in walking capacity (28). However, the presence of neurologic gait impairments in individuals post stroke can make it challenging to reach this vigorous intensity (29, 30). Locomotor high-intensity interval training (HIT) is a promising new strategy for stroke rehabilitation that uses bursts of maximum speed walking alternated with recovery periods, which allows individuals to sustain higher aerobic intensities than physiologically possible with continuous exercise (28). Adding treadmill HIT to inpatient stroke rehabilitation has been shown to significantly improve gait outcomes (31, 32). A preliminary study in chronic stroke demonstrated that treadmill HIT can elicit significant increases in walking capacity, gait speed and aerobic fitness in just 4 weeks (33). A subsequent report showed the feasibility of combining treadmill and overground HIT in an effort to better translate treadmill gait improvements into the normal overground walking environment (34). Thus, HIT serves as a promising new strategy to target impairments in aerobic fitness and motor impairment through its ability to achieve higher aerobic intensities and demonstrate improvements in walking capacity in shorter training durations.

Despite promising preliminary evidence, no previous studies have compared HIT with the current model recommended by stroke rehabilitation guidelines (MAT). In addition, the optimal training duration dose for HIT remains unknown. The present study intends to fill these gaps through completion of the following objectives: 1) Determine the optimal locomotor training intensity for eliciting immediate improvements in walking capacity among chronic stroke survivors, 2) Determine the minimum locomotor training duration required to maximize immediate improvements in walking capacity in chronic stroke, and 3) Understand the feasibility of implementing HIT at multiple sites across the United States. The primary study hypothesis is that 4 weeks of HIT will elicit significantly greater improvement in walking capacity compared to 4 weeks of MAT. Based on data from a different gait intervention in a similar population (35), we also hypothesize that compared with 4 and 8 weeks of HIT, 12 weeks of HIT will elicit significantly greater improvements in walking capacity and increased benefit over MAT.

#### Trial Design

This is a single-blind, 3-site randomized controlled trial in which participants are randomly assigned to one of two groups: locomotor moderate-intensity aerobic training (MAT) or locomotor high-intensity interval training (HIT). Prior to randomization, participants undergo a screening assessment and pre-training (PRE) blinded outcome testing to determine eligibility. Once deemed eligible, participants begin the intervention period of the study. The goal of the intervention period is to complete 36 training sessions within 12 weeks, with up to one additional week for makeup sessions in each 4-week training block. The intervention period consists of three intervention blocks separated by repeated outcome testing after 4 weeks, 8 weeks and 12 weeks of training (see Figure 1). Outcome testing is conducted by a blinded physical therapist at each time point. The primary outcome for this study is walking capacity (6-Minute Walk Test, (6MWT)), and the secondary outcomes are comfortable and fastest gait speed (10-Meter Walk Test, (10MWT)), aerobic fitness (VO_2_ at ventilatory threshold), and PROMIS Fatigue Scale total score. Exploratory measures are scores on the Activities-Specific Balance Confidence Scale, EuroQOL-5D-5L, Functional Ambulation Category, participant ratings of change, daily walking activity, spatiotemporal measures of comfortable speed instrumented walkway gait testing, resting heart rate and blood pressure, body mass index, metabolic cost of gait during treadmill exercise testing, heart rate cost of gait during 6MWT (average heart rate divided by average speed in meters/minute), difference in gait speed from the beginning to end of the 6MWT, and other measures of aerobic fitness (e.g. VO_2_ peak).

**Figure 1.**
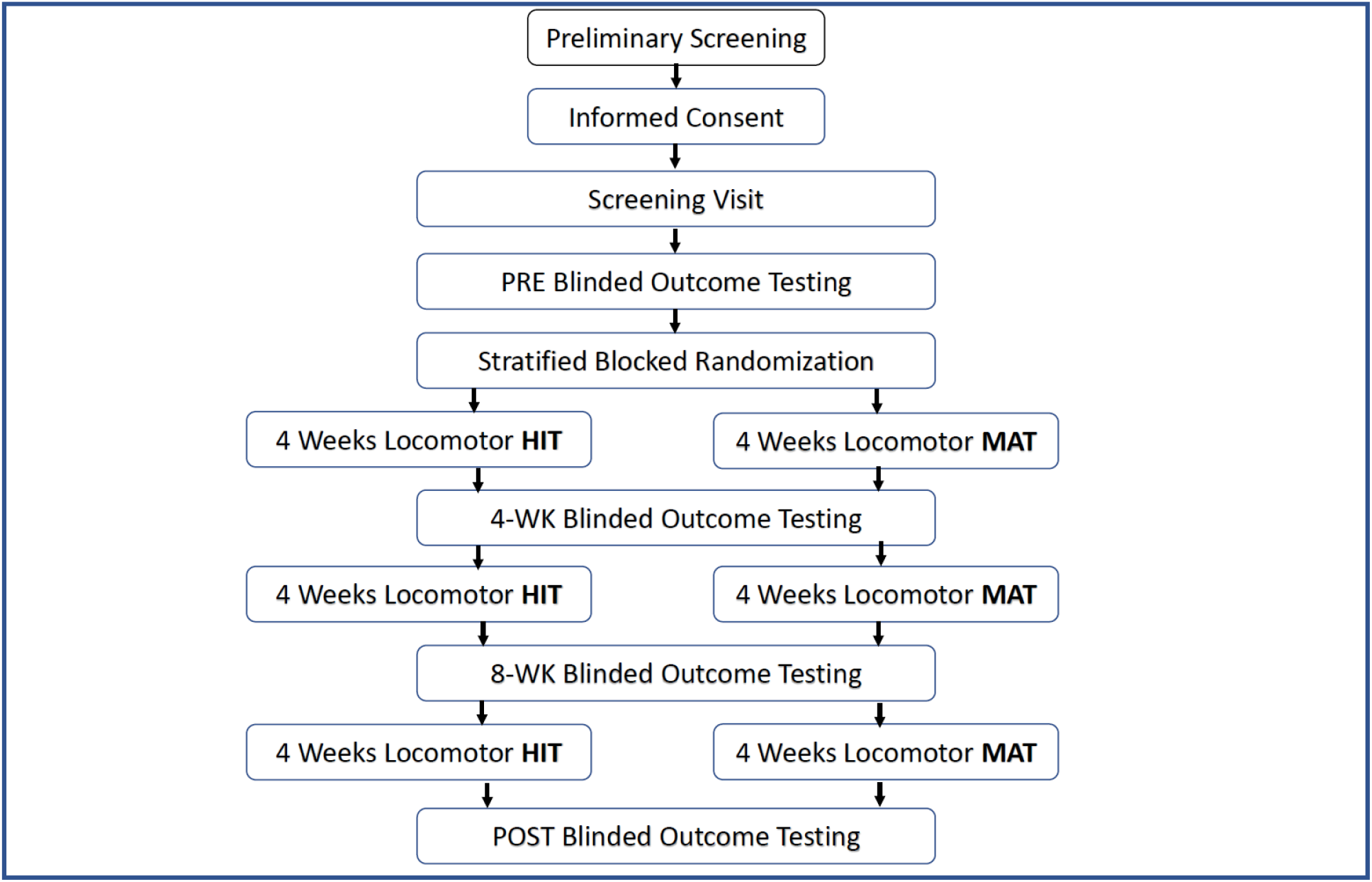
Study Schedule and Participant Flow Diagram. HIT- high-intensity interval training, MAT- moderate-intensity aerobic training

## METHODS: PARTICIPANTS, OUTCOMES, INTERVENTIONS

### Study Setting

This is a multisite clinical trial in which participants are recruited at three sites: University of Cincinnati (UC), University of Delaware (UD), and University of Kansas Medical Center (KUMC).

### Study Enrollment

The target enrollment for this study is a total of 50 participants over 3 years (approximately 6 enrolled participants per site per year). As we anticipate a screen failure rate of up to 40%, we expect to consent up to 70 participants (approximately 8 consented participants per site per year) to meet the target enrollment.

### Recruitment

Recruitment will utilize multiple approaches, including: 1) Continuous outreach to regional therapists and physicians, 2) Outreach to stroke support groups, 3) Advertisements in newspapers, magazines, social media, physician offices and/or therapy clinics, 4) Leveraging existing databases of local stroke survivors interested in participating in research, 5) Screening medical records for potentially eligible participants (UC and KUMC sites only).

### Screening Process

A member of the study team provides an overview of the study and determines initial interest in participation either in-person or via phone. The potential risks and benefits of participation are described. Potential participants are informed that participating in the study is completely voluntary and that he/she may discontinue participation at any time. For individuals who express interest, the study team member asks pre-screening questions to determine initial eligibility and answer any participant questions. For individuals who meet criteria according to pre-screening questions, a screening visit is scheduled. Each study site maintains a log to document the potential number of participants (or caregivers) contacted.

### Informed Consent

A study team member describes the study procedures and potential risks and benefits in detail at the start of the screening visit. Prior to signing informing consent, participants are asked standardized questions to ensure that the individual understands the study before consenting.

### Eligibility Criteria

The inclusion criteria for this study are as follows: 1) Age 40-80 years at the time of consent, 2) Single stroke for which the participant sought treatment, 6 months to 5 years prior to consent, 3) Walking speed ≤ 1.0 m/s on the 10-meter walk test, 4) Able to walk 10m over ground with assistive devices as needed and no continuous physical assistance from another person, 5) Able to walk at least 3 minutes on the treadmill at ≥ 0.13 m/s (0.3 mph), 6) Stable cardiovascular condition (American Heart Association class B, allowing for aerobic capacity <6 METs), 7) Able to communicate with investigators, follow a 2-step command and correctly answer consent comprehension questions.

Exclusion criteria for this study are: 1) Exercise testing uninterpretable for ischemia or arrhythmia, 2) Evidence of significant arrhythmia or myocardial ischemia on treadmill ECG graded exercise test in the absence of recent (past year) more definitive clinical testing with negative result, 3) Hospitalization for cardiac or pulmonary disease within the past 3 months, 4) Implanted pacemaker or defibrillator, 5) Significant ataxia or neglect (score of 2 on NIH stroke scale item 7 or 11), 6) Severe lower limb spasticity (Ashworth >2), 7) Recent history (<3 months) of illicit drug or alcohol abuse or significant mental illness, 8) Major post-stroke depression (Patient Health Questionnaire (PHQ-9) ≥ 10) in the absence of depression management by a health care provider, 9) Currently participating in physical therapy or another interventional study, 10) Recent botulinum toxin injection to the paretic lower limb (<3 months) or planning to have lower limb botulinum toxin injection in the next 4 months, 11) Foot drop or lower limb joint instability without adequate stabilizing device, as assessed by a physical therapist, 12) Clinically significant neurologic disorder other than stroke or unable to walk outside the home prior to stroke, 13) Other significant medical condition likely to limit improvement or jeopardize safety as assessed by a physical therapist, 14) Pregnancy, 15) Previous exposure to fast treadmill walking (>3 cumulative hours) during clinical or research therapy in the past year.

### Participant Timeline

After obtaining informed consent, participants undergo a Screening Visit to determine preliminary eligibility and record some clinical characteristics. If the participant is potentially eligible after completing the Screening Visit, the participant then performs a PRE testing visit with a blinded testing therapist to complete the eligibility assessment and obtain baseline data for the outcome measures. In total, there are four evaluation periods throughout study participation. Participants undergo evaluation assessments by a blinded testing therapist before starting the intervention period (PRE), after 4 weeks of training (4-WK), after 8 weeks of training (8-WK), and after completing 12 weeks of training (POST), see Figure 1. The evaluation procedures occurring at each of these four time points are the same and include the following measures: 6-Minute Walk Test, comfortable and fastest gait speed testing, treadmill graded exercise testing (GXT), questionnaires and recording of daily stepping activity. In addition, at the 4-WK, 8-WK, and POST evaluation time points, a global rating of change questionnaire is included that inquires about perceived changes in the participant’s walking abilities and fatigue levels as they progress through the study protocol.

### Screening Visit Eligibility Measures

During the Screening Visit, a study team member obtains the participant’s medical history, screens for the presence of depressive symptoms, performs impairment and mobility assessments of the participant, and performs StepWatch calibration procedures to begin step activity monitoring. Medical history information includes characteristics of the participant’s stroke, comorbidities, medications, surgical history, previous therapies, pain, and mobility status prior to their stroke. Additionally, the study team member instructs the participant to complete the Patient Health Questionnaire-9 to screen for depressive symptoms (36). Assistance is provided to the participant by the study team member if needed to complete the questionnaire.

The physical assessment portion of the Screening Visit consists of impairment testing and mobility assessments. During impairment testing, the following measures are administered:

- **Two-step command following:** The participant is asked to “close your eyes and make a fist”. If the individual is unable to follow two-step commands, they are ineligible for this study.
- **Lower extremity Fugl-Meyer (LEFM):** The LEFM is a stroke-specific measure of lower extremity motor impairment and includes an assessment of the participant’s reflexes and their ability to perform movements requiring different amounts of fractionated control (37). It will be used to characterize the study sample.
- **Ashworth Hypertonia Assessment:** This includes an assessment of the participant’s passive resistance to joint movement at their paretic lower extremity (38). The Ashworth Scale is scored as “excessive” (Ashworth ≥ 3) or “acceptable” (Ashworth <3). Individuals with Ashworth scores of ≥ 3 at their paretic lower extremity are ineligible for this study.
- **Ataxia and neglect testing:** Two items from the National Institutes of Health Stroke Scale (NIHSS) are used to screen for severe ataxia or neglect to determine participant eligibility (39). An assessment of upper and lower extremity coordination is used to determine the presence or absence of limb ataxia. Participants with ataxia in more than one out of four limbs (i.e. ataxia score of 2) are ineligible for study participation. Neglect testing includes an assessment of extinction to bilateral simultaneous visual and tactile stimulation. Participants who present with extinction to both sensory modalities or demonstrate behavioral evidence of profound hemi-inattention (i.e. extinction score of 2) are ineligible for study participation.

The mobility assessment of the Screening Visit consists of the following:

- **Walking-related pain assessment:** A study team member queries the participant about any pain related to walking. If the participant’s walking is limited by pain, the participant is asked further questions related to how the pain changes during walking. Based on the severity and characteristics of the participant’s pain, this may exclude them from study participation if it is determined that pain is likely to limit improvement or jeopardize safety.
- **Comfortable and fast gait speed measurements:** The 10MWT is used to assess the participant’s comfortable and fastest gait speeds. An untimed 2-meter acceleration distance is provided, followed by 10 meters of timed walking, and a 2-meter untimed deceleration distance. Participants are asked to use the assistive and orthotic devices that they most often use when walking. During the Screening Visit, comfortable gait speed is averaged over two trials. Participants whose average gait speed is greater than 1.0 m/s are excluded from study participation. If eligibility criteria are met at comfortable gait speed, a 10MWT at the participant’s fastest gait speed is performed. The 10MWT is a valid and reliable measure of walking speed in individuals post stroke (40).
- **Height and weight assessment:** The participant’s height and weight are obtained at the Screening Visit. Weight is reassessed at the 4-WK, 8-WK, and POST time points and used to normalize metabolic data from GXT testing at these time points.
- **Treadmill acclimation and screening assessment:** This assessment is used to determine eligibility and for preparation for exercise testing. As this study involves considerable treadmill walking, the treadmill acclimation provides the participant with the opportunity to acclimate to the treadmill and provides the study team with an idea of how fast the participant can safely walk on the treadmill. During treadmill walking, the participant’s heart rate (HR) is monitored using a Polar heart rate monitor synched to the Digifit iCardio app to provide real-time HR monitoring. The study team member administering the test ensures that the participant’s HR does not exceed 80% age-predicted heart rate reserve (HRR) during this test. The participant wears a harness that is attached to an overhead system for protection in the event of a fall and is asked to hold onto the handrail during testing. The treadmill is started at a slow speed and gradually increased in communication with the participant. The speed goal for this test is the participant’s fast overground speed or 0.3 mph, whichever is higher. Rest breaks are provided to maintain <80% HRR, and a post-exercise blood pressure is taken upon completing the test. Participants must be able to walk at least 0.3 mph on the treadmill to be eligible for study participation.

After the Screening Visit (but before the PRE evaluation visit), pertinent medical records are obtained to ensure the participant is safe to participate in the study. A radiology report confirming stroke and its location is also obtained. Participants who do not meet one or more of the eligibility criteria that can be assessed from the Screening Visit are excluded (see Eligibility Criteria above).

### Step Activity Monitoring

For participants that meet eligibility criteria at the time of the Screening Visit, a step activity monitor (Modus StepWatch) is calibrated and issued to the participant to wear throughout the study protocol during all waking hours except bathing. The StepWatch is calibrated per the manufacturer’s instructions and synced to an iPad app that enables study team members to download the participant’s stepping activity throughout the study protocol. At the start of each subsequent study visit, a study team member “reads” the step activity data and records the stepping activity and respective dates in the study’s electronic data management system, REDCap (Research Electronic Data Capture). Stepping activity data is also recorded at the end of each study visit which enables study team members to discern steps taken within each study session from steps taken outside of study visits. While reading step activity data, the study team member checks for data irregularities (e.g. missing steps in the first or last half of non-study-visit days) and queries the participant to determine if a particular day should be deemed a valid recording day. If it is likely that >10% of walking bouts that day were not recorded, then it is documented as not a valid recording day.

### Outcomes

The following measures are assessed at the PRE, 4-WK, 8-WK, and POST evaluation time points and are conducted by a licensed physical therapist who is blinded to group randomization. The goal is to complete all of the tests in the same visit and within 2-7 days since the last training session in the preceding intervention block. Whenever possible, gait testing (i.e. 6MWT and 10MWT) is completed prior to graded exercise testing, as gait testing includes the primary outcome measure. Blood pressure and HR are taken at each visit. For all questionnaires, the participant is encouraged to self-administer (if able) to reduce any influence that the study team member may have on their responses.

- **6-Minute Walk Test (6MWT):** The 6MWT is a measure of walking capacity and is the primary outcome measure for this study. Study team members ensure that the participant is provided with adequate rest prior to performing this measure. Participants are instructed to walk as far and fast as they can for six minutes and are asked to use the orthotic and assistive devices they most often use during normal daily walking (41). Participants walk along a marked pathway that was required to be at least 20 meters long at each site. At UC and KUMC, participants walk back and forth between two cones spaced 25 meters apart. At UD, participants walk around a rectangular course with a 103.6-meter perimeter. The participant is informed that they may stop and rest as needed but that the timer will keep counting down. Participants are notified how much time has elapsed in 1-minute intervals, and HR is monitored and continuously recorded throughout the test. The total distance walked, time to walk the first 25-meter length or 100 feet, time to walk the last complete 25-meter length or 100 feet, average HR, and max HR are recorded in REDCap. The 6MWT is a valid and reliable measure of walking endurance in individuals post stroke (40, 42).
- **Comfortable and Fast 10-meter Walk Test:** The participant’s comfortable and fast gait speeds are assessed using the 10MWT in the same manner as described above. Two comfortable speed trials and two fast speed trials are taken at each of the four outcome testing time points.
- **Treadmill Exercise Testing with Metabolic Cart:** At each evaluation time point during the study, a treadmill graded exercise test (GXT) is performed. Exercise testing is performed on a motorized treadmill with a 12-lead electrocardiogram (ECG) and metabolic cart for analysis of VO_2_. At the start of the test, the participant’s resting vital signs and ECG are obtained. Participants then walk on a treadmill wearing a harness attached to an overhead system for fall protection and hold onto the treadmill handrail. The starting treadmill speed is 0.3 mph for the first 3 minutes and then gradually increases in increments of 0.1 mph every 30 seconds until peak volitional exertion. The incline of the treadmill remains 0% unless the participant achieves a speed of 3.5 mph at which time the incline increases in increments of 0.5% every 30 seconds while the speed remains fixed. Ratings of perceived exertion (RPE) (43) and blood pressure are assessed every two minutes during the test. Test termination criteria include the participant’s request to stop, the participant drifting backward on the treadmill and being unable to recover, gait instability judged to pose an imminent safety risk by the testing therapist, and other stop criteria according to American College of Sports Medicine guidelines (44). After completion of this test and a 10-minute rest period, participants are asked to attempt a 3-minute verification test to determine whether maximum heart rate was reached. During the verification test, the speed is increased to the last successfully completed stage from the GXT, and a 3-minute timer is started once the treadmill has reached this target stage. Participants are encouraged to try and complete the full 3 minutes of the verification test when possible. VO_2_ is not measured during the verification test. Guidelines for stopping criteria are the same as the GXT, except that the verification phase can be stopped once the participants completes the 3 minutes. The instantaneous peak HR from the verification test is recorded in the study’s electronic database. During the PRE period, a physician or medical monitor reviews the test results to ensure safety to continue with the study protocol. In this study, the peak heart rate achieved during the GXT or verification test (whichever is higher) is used to derive training intensity zones. If the participant does not achieve 85% of their age-predicted maximum heart rate during the GXT or verification test (44), the peak GXT or verification test heart rate (whichever is higher) is also used as a heart rate limit during subsequent training sessions. VO_2_ at the ventilatory threshold is a secondary outcome for this study as evidence suggests that this measure may be a more valid assessment of aerobic capacity compared to VO_2_ peak in individuals with stroke (45).
- **PROMIS Fatigue Scale (Version 8a):** The PROMIS Fatigue Scale is an 8-item self-report questionnaire that inquires about the participant’s fatigue over the past seven days (46). Responses are rated on a 5-point Likert scale from ‘not at all’ to ‘very much’.
- **Functional Ambulation Category (FAC):** The FAC is a measure of walking independence and is scored on a scale of 0-5 (47). A member of the study team rates the participant’s level of walking independence based on their walking performance during the 10MWT and 6MWT. For this study, only scores of 2-4 are permitted as individuals obtaining lower scores would not meet the eligibility criteria, and a score of 5 would require observation of the participant walking on different types of non-level surfaces, which is not part of this study.
- **EQ-5D Quality of Life Questionnaire (Version 5L):** The EQ-5D is a 6-item questionnaire about quality of life as it relates to mobility, self-care, usual activities, pain/discomfort, anxiety/depression, and overall health (48).
- **Activities Specific Balance Confidence Scale (ABC):** The ABC is a 16-tem questionnaire that asks participants to rate their balance confidence during everyday tasks on a scale from 0% to 100% (49). Scores are averaged to provide an overall value representing the participant’s perceived balance confidence.
- **Global Rating of Change (GROC):** This questionnaire asks participants to rate their perceived change in areas related to their walking abilities and walking habits since beginning the study (not applicable at PRE testing) (50). Responses are scored on a 7-point ordinal scale ranging from ‘much better’ to ‘much worse’.
- **Electronic Walkway Gait Assessment:** Before starting the first training session in each intervention block (i.e. training sessions 1, 13, and 25) and the final training session (i.e. session 36), the participant’s comfortable speed gait parameters are recorded with two passes across an electronic walkway (e.g. GaitRITE). Participants are asked to use their habitual assistive and orthotic devices. The following gait parameters are recorded in REDCap: gait velocity, cadence, right and left step lengths (cm), right and left step times (s), right and left single limb support (% of gait cycle), and right and left stride velocity (cm/s).

### Allocation

Eligible participants are randomized after the PRE blinded outcome testing visit and before the first training session. Participants are randomized in a 1:1 ratio to either HIT or MAT, using the REDCap randomization module. This module ensures concealed allocation by requiring the study team member to confirm participant eligibility prior to revealing the randomization allocation and not permitting anyone to un-randomize a participant. The study statistician who computer-generated the randomization sequence and uploaded it to REDCap is the only person who has access to view it and has no interaction with study participants (e.g. not involved with recruitment or enrollment). Randomization is stratified by site and baseline walking speed (<0.4, ≥ 0.4 m/s) to help ensure that groups are balanced within sites and on this critical prognostic factor (51, 52). Within each stratum, block size is randomly permuted to prevent study personnel from being able to predict the last randomization within a block. SAS® (SAS Institute, Cary, NC) PROC PLAN was used to create the randomization scheme.

### Interventions

Participants are randomly assigned to either MAT or HIT. Study interventions are administered under the direction of a licensed physical therapist. For each intervention block, the goal is to complete 12 training sessions within 4 weeks, with an additional week allowed for makeup sessions. The target frequency of training is 3 sessions per week (with one day of rest between training sessions, when possible).

The following procedures are common to both intervention groups. Each training visit involves 45 minutes of exercise that consists of a 3-minute warm-up of overground walking, 10 minutes of overground training, 20 minutes of treadmill training, 10 minutes of overground training, and a 2-minute cool-down of overground walking. Throughout training, participants use their customary orthotic devices. During overground training, participants use the assistive device that best enables achievement of intervention goals (fastest speed for HIT; target HR for MAT). The participant’s overground gait training speed is measured at the beginning and end of each overground bout. During treadmill walking, participants wear a harness connected to an overhead support system for fall protection and are asked to use a handrail for balance support. Guarding is provided by the training therapist to help prevent falls or injury. No assistance or cueing is provided to improve the participant’s gait pattern.

During training, participants wear a heart rate monitor and step activity monitor to monitor heart rate and stepping activity, respectively. Heart rate is monitored using the Polar H7 Bluetooth 4.0 transmitter synched to an Apple iPod application (Digifit iCardio) to enable continuous HR monitoring throughout the training session. The iPod application is also used to time the duration of each component of training (e.g. 20-minute treadmill bout). The target HR for training sessions is based on the participant’s highest HR achieved during the GXT or verification test. Resting HR values are obtained at the start of the training visit in a standing position. Stepping activity data is recorded before and after each treatment session to monitor steps taken during the session.

Rating of Perceived Exertion (RPE), blood pressure, and blood lactate are also monitored throughout the intervention protocol. Participants are shown an RPE chart at the end of each training session and asked how hard they were working during the session on average. Blood pressure is monitored at least once per session until a consistent response within safety limits is established. Blood lactate concentrations are measured in the middle session of each training week (i.e. every 3 training sessions starting at session 2) immediately after completing the treadmill training portion of the session. Immediately after treadmill training, the participant is instructed to sit on a chair on the treadmill, and the training therapist obtains a measure of blood lactate via fingerstick with caution taken to avoid sweat contamination and alterations in lactate concentration due to vigorous finger squeezing.

If any of the following occur during a training session, exercise is paused (timer will continue) and the training therapist decides whether early termination and/or physician notification is warranted: 1) New onset pain, 2) HR consistently exceeding peak HR achieved on most recent GXT or verification test (only if the participant has not reached 85% age-predicted maximum heart rate during exercise test), 3) Difficulty monitoring heart rate or blood pressure, 4) Participant requests a break. If any of the following occur during a training session, the session is terminated, the participant’s physician is notified, and the site primary investigator decides whether to withdraw the participant from the study: 1) Signs of poor perfusion, 2) Drop in systolic blood pressure ≥ 10 mmHg below the resting level from that day despite an increase in workload, 3) Hypertensive response with systolic blood pressure >240 mmHg and diastolic blood pressure >110 mmHg, 4) New onset of significant nervous system symptoms or claudication pain, 5) Chest pain or angina, 6) Severe fatigue or shortness of breath in excess of what would be expected from exercise, 7) Serious injury.

#### Locomotor moderate-intensity aerobic training (MAT)

Individuals randomized to the MAT group perform continuous walking on the treadmill and overground. During training, speed is continuously adjusted to maintain the following target HR ranges: Training Sessions 1-6: 40 ± 5% HRR; Training Sessions 7-12: 45 ± 5% HRR, Training Sessions 13-18: 50 ± 5% HRR; Training Sessions 19-36: 55 ± 5% HRR. All attempts are made to keep heart rates below 60% HRR during MAT training sessions as this is generally considered the threshold for vigorous intensity (44). During overground MAT, the participant is instructed to walk continuously for 10 minutes, and the training therapist instructs the participant to speed up or slow down to maintain their heart rate in the desired training zone.

For treadmill MAT, participants walk continuously for 20 minutes if possible. At the start of each session, the training therapist selects a speed that brings the participant as close as possible to, but not exceeding, the target HR. For the first training session, treadmill speeds start at ∼75% of the participant’s comfortable gait speed from the most recent 10MWT. The training therapist then adjusts the speed as needed to keep the participant’s HR in the target zone. The training therapist decreases the speed if the participant requests a speed decrease, the participant drifts backward and does not immediately recover, gait instability is observed and judged to pose an immediate safety risk, toe drag that persists into mid-swing is observed, or there is evidence of excessive joint instability with risk of harm.

#### Locomotor high-intensity interval training (HIT)

Individuals randomized to the HIT group perform repeated 30 second bursts of walking at their maximum safe speed, alternated with 30-60 second rest periods. During overground HIT, burst speed is increased using visual feedback about the distance covered during each burst and encouragement to increase distance. During treadmill HIT, speed is systematically increased throughout each training session based on performance criteria. Speed is the primary intensity target for the HIT group, and HR is secondary after speed is maximized. This is primarily because the 30 second bursts are not long enough for heart rate to reach steady state, so it fluctuates between burst and recovery and trends upward over the session (53, 54). The target average HR for each session is ∼70% HRR, with a range from 60% to 95% HRR. If the participant reached their target HR of 85% of their age-predicted maximum (not adjusted for beta-blockers) during the GXT and had normal results, then no HR limit is enforced. However, if the participant did not reach their target HR once during any previous GXTs, their training HR is limited to their maximum HR achieved across all previous GXTs.

During overground HIT training, the participant is instructed to walk as fast as they can for 30 seconds. A marker is placed at the participant’s starting position as well as their final position after the 30-second burst. For future bursts, participants are encouraged to achieve at least the distance they covered during previous bursts and further if they are able. Sixty seconds of rest is provided after the first three bursts and then decreased to 30 seconds rest periods thereafter. However, the training therapist may consider extending the rest periods if the participant needs to sit down during recovery, if the distance covered during the previous bursts significantly decreases with shorter rest periods, if the participant requests an extended rest break, or if the participant exceeds their heart rate limit.

When selecting speeds for treadmill HIT during bursts, the goal is to quickly find the participant’s fastest safe challenge speed and increase this speed as able throughout the burst. The challenge speed is defined as the speed at which the participant can safely complete the burst but has some backward drift or gait instability with recovery. During the first treadmill HIT session, treadmill speeds start at ∼75% of the participant’s peak successful speed from their most recent GXT. To determine an initial challenge speed during bursts, the training therapist waits 15 seconds to allow the speed to ramp up and the participate to acclimate, and then increases the speed by 0.1 mph every 5 seconds. Once the challenge speed is found, specific criteria are used to determine whether subsequent burst speeds will be increased, maintained, or decreased. If a burst is performed safely with no gait instability or backward drift, the speed is increased by 0.1 mph for the next burst. If the challenge speed criteria are met, the speed is kept the same for the next burst. If a burst is not performed safely or must be stopped early due to backward drift without recovery or unsafe gait instability, the speed is decreased by 0.1 mph for the next burst. Similar to overground HIT, 60 seconds of rest are provided in between the first three bursts, followed by 30 second rest periods between subsequent bursts and similar criteria are used during treadmill HIT to determine if an extended rest period is required.

### Personnel Training and Standardization

A systematic training and competency assessment program for all study therapists and coordinators has been implemented to maintain standardization of study procedures across sites. Study personnel cannot perform an official study role until certified for that role. The site primary investigator (PI) and site coordinator ensure that the study team member meets competency requirements. Study personnel training procedures include the following: 1) Reading the study manual of operating procedures (MOP), 2) Complete online personnel training modules related to their study roles 3) Practice using the study’s electronic data management system, REDCap, 4) Practice using all equipment required for their study roles. Trainees must show competency in all aspects of their role before certification. Recertification is done as needed based on the discretion of the site PI. A delegation of authority log is maintained at each site to delineate the job roles of study team members. Communication between the site PIs is maintained through meetings, conference calls, or emails as needed to maintain consistency in study procedures across sites.

## METHODS: DATA MANAGEMENT AND ANALYSIS

### Data Management

This study uses both electronic and hard-copy data management procedures. For electronic database management, the secure data platform REDCap is used, and the majority of study data is directly entered into the REDCap database using an iPad during each study visit. This database includes automated calculations, quality control checks and prompts (e.g. notification if entered data indicate participant is not eligible, calculation of intensity targets, notification of whether entered intensity data are within target range, prompts to fill in any missing data or to double check any values outside of the expected range). Each study team member is provided a secure login for the University of Cincinnati REDCap portal through the UC regulatory coordinator and provided data access rights based on their study role such that blinded personnel cannot access randomization or intervention data.

This study also generates some electronic data outside of REDCap that could be further processed to obtain additional variables of interest. Examples of such data include metabolic cart files, electronic walkway files, and StepWatch activity files. These data files are uploaded and stored in a secure OneDrive folder that is designated for research data so that they can be processed centrally. To maximize data security, no participant identifiable information is entered into these files or the software that creates these files. In addition, hard-copy records containing information that could be used to identify participants (e.g. consent forms, medical records) and any hard copy forms containing study data (e.g. temporary backup paper forms in case of power, internet or REDCap server downtime) are maintained in a locked storage unit inside a controlled-access room throughout the study. Regulatory documents are maintained according to institutional requirements and guidelines specific to each site.

### Sample Size

This study is powered to detect the minimally clinically important difference (MCID) of 20 meters in walking capacity (6MWT) change between groups (55). The 6MWT change estimate for the MAT group was extrapolated from a 4-week pilot study and resulted in a change estimate of 15 meters every 4 weeks (33). The 6MWT change estimate for the HIT group was calculated by adding the MCID to the MAT group estimate (15+20=35 meters every 4 weeks). Variance and covariance parameters were estimated by pooling data across two previous 4-week studies (n=20), using the mean variance for each time point and the highest suggested exponential decay rate (0.5) (56) for the repeated measures correlations to extrapolate parameters for the 8-WK and POST time points. These calculations indicated a target sample size of 40 (20/group) for 80% power. To account for up to 20% attrition, the target enrollment is 50 participants.

### Statistical Methods

SAS v9.4 will be used for data analysis, and the study statistician will remain blinded to study group. Data related to baseline variables, intervention fidelity and concurrent outside interventions will be compared between groups using t-tests and X^2^. If a baseline prognostic factor is found to differ between groups, it will be considered for inclusion as a covariate during hypothesis testing. The primary analysis will follow intent-to-treat methods and any missing data will be handled with the maximum likelihood method, assuming that patterns of missingness do not violate the missing at random assumption (57). To test robustness of different ways to handle missing data, sensitivity analyses will be used.

#### Hypothesis 1

To test our primary hypothesis that, compared with 4 weeks of MAT, 4 weeks of HIT will elicit significantly greater improvement in the 6MWT distance, a general linear model will be used. In this model, we will use fixed effects for group (HIT, MAT), time (PRE, 4-WK, 8-WK, POST), [group x time], site (UC, KUMC, UD), [site x time], baseline speed category (<0.4, ≥0.4 m/s), and [baseline speed category x time] with an unstructured covariance matrix. This hypothesis will be tested by the significance of the [group x time] contrast from the PRE to 4-WK for the 6MWT at α=0.05. Secondary outcomes will be tested separately using this same model to identify the most sensitive measures to carry forward into future studies (58). The Benjamini-Hochberg procedure (59) will be used to control the false discovery rate for the secondary outcomes.

#### Hypothesis 2

To test the hypothesis that, compared with 4 and 8 weeks of HIT, 12 weeks of HIT will elicit significantly greater improvements in walking capacity and increased benefit over MAT, the same general linear model described above will be used. The hypothesis that 12 weeks of HIT will elicit greater improvements in primary and secondary outcomes compared to 4 and 8 weeks of HIT will be tested by the significance of the respective time contrasts within the HIT group. The hypothesis that HIT will elicit significantly greater improvements in primary and secondary outcomes from PRE to 8-WK and PRE to POST compared to MAT will be tested by the significance of the respective [group x time] contrasts. False discovery rate control will be applied for secondary outcomes (59).

We will also test for baseline cofactors that may influence a stroke survivor’s response to the interventions in this study. To do this, we will utilize a multivariate prognostic model that includes comfortable gait speed, lower extremity Fugl-Meyer motor scores, and scores on the Activities-Specific Balance Confidence Scale. These measures were selected based on previous studies suggesting that comfortable gait speed (52, 60-64), lower limb Fugl-Meyer motor scores (64-66), and balance abilities (67) may influence response to gait rehabilitation interventions in individuals with chronic stroke. Other potential cofactors will also be explored to inform future studies.

Based on safety data from preliminary studies (33, 53) and extensive previous HIT research among participants with heart disease (24, 68-74) and MAT research among individuals post stroke (15), we expect a similar rate of non-serious adverse events (AEs) between HIT and MAT (e.g. temporary exercise-related soreness and fatigue), without any study-related serious AEs. In the unexpected event of one or more serious adverse events (SAE), the SAE rate will be compared between groups to confirm that there is no significant difference in major safety risk between HIT and MAT. A logistic regression model will be used for this analysis with SAE (yes/no) as the dependent variable and fixed effects for group, site, and baseline gait speed category. If there are SAE(s) in one group only, a continuity correction (0.5 SAEs added to each group) will still allow the odds ratio to be calculated (33).

## METHODS: MONITORING

### Data Monitoring

The Data and Safety Monitoring Board (DSMB) for this study consists of three independent members separate from the study team at institutions outside of UC, UD and KUMC. Collectively, the DSMB has experience in the management of patients with stroke, exercise, and clinical trials. Persons with a significant conflict of interest were not permitted to be DSMB members. The role of the DSMB is to monitor participant accrual, randomization balance and safety data to assess the risks of study participation.

The DSMB meets annually throughout the study, either in person or via teleconference. Additional meetings may be scheduled as requested by the investigators, IRB or DSMB members. The DSMB remains blinded unless it requires the group identities to perform its duties. DSMB meetings include open sessions where the DSMB may discuss any issues with the study team as well as closed sessions where the DSMB alone decides on its recommendations. After each DSMB meeting, the DSMB provides a written report of their discussions and recommendations as to whether the study should continue, whether modifications to the study are needed, or if the study should be terminated. These reports are sent to the investigators, the Institutional Review Board (IRB) and the sponsor. The study may be modified or discontinued at any time by the research team, DSMB, IRB or sponsor to ensure the protection of research participants.

#### Outcome Data Monitoring

A blinded co-investigator monitors REDCap outcome data for missing or implausible values.

#### Study Intervention Fidelity Monitoring

The site PIs and/or coordinators monitor REDCap intervention data for missing or implausible values and intervention fidelity. Monitored data include the following:

- **Adherence:** This is measured by the number of training sessions attended and completed.
- **Aerobic intensity:** This includes the mean and maximum training session HR relative to the target HR range and relative to the previous training sessions. It also includes time spent in target HR zones.
- **Anaerobic intensity:** This is measured using blood lactate concentration after the treadmill training portion of one session each training week, using a finger stick and a point-of-care blood lactate analyzer.
- **Neuromotor intensity:** This is measured by treadmill and overground training speeds each session.
- **Repetition of practice:** This includes step counts during each session, measured by an activity monitor placed on the participant’s non-paretic lower extremity.

### Adverse Event and Protocol Deviation Reporting

All identified AEs and protocol deviations are reported to the UC IRB and DSMB annually. Unanticipated problems requiring prompt reporting are reported per UC IRB policy (described below). AEs and protocol deviations are reported by study staff to the site PIs on a regular basis and are discussed during study conference calls.

We define an AE as ‘any unfavorable and unintended sign, symptom, or disease temporally associated with study participation that may or may not be related to study procedures, including any adverse change that occurs at any time following consent and before completing study participation’. An SAE is an AE that results in any of the following outcomes: death, a life-threatening situation, inpatient hospitalization or prolongation of existing hospitalization, or a persistent or significant disability/incapacity. Important medical events that may not result in death, be life-threatening, or require hospitalization may also be considered SAEs when, based on appropriate medical judgment, they may jeopardize the patient and may require medical or surgical intervention to prevent one of the outcomes in this definition.

Anticipated AEs are either listed in the study protocol or consent form or have a reasonable likelihood of occurrence in the study population (adults and older adults with stroke). Possible events listed in the protocol or consent form (regardless of likelihood) include discomfort, worry, pain, fatigue, stiffness, skin breakdown, local infection, faintness, nausea, bruising, scarring, fall, injury, myocardial infarction, or other serious heart problems. Events with greater likelihood of occurrence in adults and older adults with stroke include (but are not limited to): pain, fatigue, stiffness, faintness, syncope, vertigo, fall, skin breakdown, bruising, orthopedic injury, recurrent stroke, angina, myocardial infarction, blood clot and seizure.

All identified AEs are named using terminology from the Common Terminology Criteria for Adverse Events version 4.03 (CTCAE, National Cancer Institute, 2010); however, “Dizziness” will be categorized as either “Lightheadedness” or “Vertigo”. The CTCAE criteria are also used as a guideline to provide a severity grade for the AE which will range from 1 (mild) to 5 (death). For this study, a serious adverse event is defined as grade ≥ 3/5 (75).

The relationship of an AE to study testing or interventions is determined using pre-defined criteria that considers when the event occurs in relation to testing/training procedures, whether the AE follows a pattern consistent with study procedures, whether it improves when the procedure has stopped or reappears when the procedure is resumed or repeated, and whether an alternative cause or influence may also be present. We also consider the impact of the AE on study interventions. The AE is considered to have no impact on study interventions if study interventions do not require any alteration because of the event. Modification to study interventions would occur when study interventions are modified such that they differ in a substantive way from what is described in the protocol because of the AE. The AE may also result in termination such that the participant withdraws or is withdrawn from the study before completing the intervention because of the AE.

Participants are queried about any adverse events at the start and end of each study visit. AEs that are specifically queried include falls, injuries, pain, lightheadedness and fatigue. During study visits, participants are monitored for signs or symptoms of cardiorespiratory insufficiency, new neurologic impairments or orthopedic injury. Whenever a study team member identifies an AE, an Adverse Event Form in REDCap is started and any additional information needed is collected. If the AE is not already resolved when discovered, the study team member follows up on the AE during each visit and/or by phone until it is resolved. The study team member completing the AE form provides a description of the event, its severity, its timing relative to study testing and/or intervention procedures, any possible alternative causes or contributing factors, any AE-related interventions (e.g. pain medicine), any follow up, and if/when the event is resolved. The study team member completing the form also preliminarily grades and categorizes the event using the above guidelines. All AE reports identify participants only by their study ID as these reports are viewed by blinded study team members. Once resolved, AE reports are adjudicated by the blinded study physician to determine the official severity grade and categorization using the information provided by the study team member (the blinded study physician may also request additional information if needed). Withdrawal from the study and modifications to study procedures as a result of an AE or because of therapeutic measures taken to treat an AE are at the discretion of the site PIs, in consultation with the study neurologists or cardiologists as appropriate.

A protocol deviation is when one or more procedures described in the study IRB protocol are not followed, either intentionally or unintentionally. Each site maintains a protocol deviation log that is sent to the UC regulatory coordinator upon request. This log includes: 1) a description of the protocol deviation, 2) the date of the deviation, 3) the participant ID(s) affected, 4) whether the protocol deviation was related to screening/enrollment, outcome testing, and/or study intervention, and 5) either a description of the corrective actions taken to prevent recurrence or a rationale of why such actions are not needed.

All AEs and protocol deviations are reported by study staff to site PIs on a regular basis and discussed on study conference calls. Any major AEs or protocol deviations are reported to site PIs and the UC PI as soon as possible. All identified AEs and protocol deviations are compiled by UC and reported to the IRB and DSMB annually. The IRB of record for all sites in this study is the University of Cincinnati and requires prompt reporting (within 10 days of discovery) of any unanticipated problems involving risk to participants or others. Events resulting in temporary or permanent interruption of the study activities by a site PI to avoid potential harm to participants are reported to the UC IRB within 48 hours of discovery. The lead site PI reviews the event and determine if it meets criteria for prompt reporting.

## DISCUSSION

This is the first study designed to compare HIT and MAT post-stroke and the first to compare different HIT durations. Previous work has shown that among healthy adults, HIT delivers significant benefits remarkably faster (within 6 sessions over 2 weeks) (76-78), achieving similar improvements to MAT with up to 76% less training time (77, 79-81). If HIT elicits comparable changes among individuals with stroke in 4 weeks of training (objective 1 of this study), it would provide a clinically feasible and resource-efficient alternative to the current best-practice model (MAT), which could result in increased exercise engagement among stroke survivors. In addition, no previous studies have compared different HIT durations or examined the time course of outcome changes. This study intends to fill that gap (objective 2) and will provide foundational information to guide dosing of locomotor intensity and duration in future studies and clinical practice.

This study is also the first U.S. multi-site trial of post-stroke HIT. Thus, the results of this study will also aid in our understanding of the feasibility of implementing HIT across multiple sites nationally (objective 3). Depending on the results of this research, the next step would be a larger efficacy trial. To that end, this study will provide needed data to design a subsequent definitive trial of the relative efficacy of HIT and MAT for eliciting clinically meaningful and sustained improvements in walking function.

## Data Availability

Data sharing is not applicable to this article as the datasets are currently being generated and have not yet been analyzed. De-identified data will be deposited in the National Institute of Child Health and Human Development (NICHD) Data and Specimen Hub (DASH) repository.

## Trial Status

Protocol version 2020-01-21. Recruitment start date: 2019-01-04. Estimated completion date: 2022-02-28.

## LIST OF ABBREVIATIONS

MAT: Moderate-intensity aerobic training;
HIT: High-intensity interval training;
10MWT: 10-Meter Walk Test;
6MWT: 6-Minute Walk Test;
VO_2_-peak: peak oxygen uptake;
UC: University of Cincinnati;
UD: University of Delaware;
KUMC: Kansas University Medical Center;
MOP: Manual of Operating Procedures;
RPE: Rating of Perceived Exertion;
GXT: Graded Exercise Test;
PRE: pre-blinded outcome testing;
4-WK: 4-week blinded outcome testing;
8-WK: 8-week blinded outcome testing;
12-WK: 12-week blinded outcome testing;
LEFM: Lower Extremity Fugl-Meyer;
NIHSS: National Institutes of Health Stroke Scale;
FAC: Functional Ambulation Category;
ABC: Activities Specific Balance Confidence Scale;
GROC: Global Rating of Change;
ECG: electrocardiogram;
PI: Primary Investigator;
HR: Heart Rate;
HRR: Heart Rate Reserve;
MCID: Minimally Clinically Important Difference;
DSMB: Data and Safety Monitoring Board;
AE: Adverse Event;
SAE: Serious Adverse Event;
CTCAE: Common Terminology Criteria for Adverse Events

## DECLARATIONS

### Ethics Approval and Consent to Participate

This research has been approved by the University of Cincinnati IRB. The University of Cincinnati IRB is the IRB of record for this protocol, and the agreement is managed through SMART IRB. All participants are providing written informed consent prior to participation.

### Consent for Publication

Not applicable-this manuscript does not contain an individual person’s data.

### Authors’ Contributions

AM primary author, primary testing PT at UD. DSR secondary author, UD site PI. SB author, KUMC site PI. KD author, blinded co-investigator. SD author, study coordinator at UC. JW author, KUMC site coordinator. HW author, primary training PT at UD. EW author, primary study coordinator at UC. DC author, co-investigator, blinded centralized assessor of ventilatory thresholds. MG author, co-investigator, study cardiologist. OA author, co-investigator, blinded adverse event adjudicator. JK author, co-investigator, study statistician. BK author, co-investigator, study neurologist. PB senior author, Principal Investigator. All authors read and approved the final manuscript.

## Acknowledgements

Not applicable.

## Authors’ Information (optional)

Not applicable.

## References

1. Mozaffarian D, Benjamin EJ, Go AS, Arnett DK, Blaha MJ, Cushman M, et al. Heart Disease and Stroke Statistics-2016 Update: A Report From the American Heart Association. Circulation. 2016;133(4):e38–360.

2. Winstein CJ, Stein J, Arena R, Bates B, Cherney LR, Cramer SC, et al. Guidelines for Adult Stroke Rehabilitation and Recovery: A Guideline for Healthcare Professionals From the American Heart Association/American Stroke Association. Stroke. 2016;47(6):e98–e169.

3. Ada L, Dean CM, Lindley R, Lloyd G. Improving community ambulation after stroke: the AMBULATE Trial. BMC Neurol. 2009;9:8.

4. Hill K, Ellis P, Bernhardt J, Maggs P, Hull S. Balance and mobility outcomes for stroke patients: a comprehensive audit. Aust J Physiother. 1997;43(3):173–80.

5. Jorgensen HS, Nakayama H, Raaschou HO, Vive-Larsen J, Stoier M, Olsen TS. Outcome and time course of recovery in stroke. Part II: Time course of recovery. The Copenhagen Stroke Study. Archives of physical medicine and rehabilitation. 1995;76(5):406–12.

6. Mayo NE, Wood-Dauphinee S, Cote R, Durcan L, Carlton J. Activity, participation, and quality of life 6 months poststroke. Archives of physical medicine and rehabilitation. 2002;83(8):1035–42.

7. Bohannon RW, Horton MG, Wikholm JB. Importance of four variables of walking to patients with stroke. Int J Rehabil Res. 1991;14(3):246–50.

8. Billinger SA, Arena R, Bernhardt J, Eng JJ, Franklin BA, Johnson CM, et al. Physical activity and exercise recommendations for stroke survivors: a statement for healthcare professionals from the American Heart Association/American Stroke Association. Stroke. 2014;45(8):2532–53.

9. Marsden DL, Dunn A, Callister R, Levi CR, Spratt NJ. Characteristics of exercise training interventions to improve cardiorespiratory fitness after stroke: a systematic review with meta-analysis. Neurorehabil Neural Repair. 2013;27(9):775–88.

10. Pang MY, Charlesworth SA, Lau RW, Chung RC. Using aerobic exercise to improve health outcomes and quality of life in stroke: evidence-based exercise prescription recommendations. Cerebrovasc Dis. 2013;35(1):7–22.

11. Stoller O, de Bruin ED, Knols RH, Hunt KJ. Effects of cardiovascular exercise early after stroke: systematic review and meta-analysis. BMC Neurol. 2012;12:45.

12. Kendall BJ, Gothe NP. Effect of Aerobic Exercise Interventions on Mobility among Stroke Patients: A Systematic Review. American journal of physical medicine & rehabilitation. 2016;95(3):214–24.

13. Mehta S, Pereira S, Janzen S, Mays R, Viana R, Lobo L, et al. Cardiovascular conditioning for comfortable gait speed and total distance walked during the chronic stage of stroke: a meta-analysis. Topics in stroke rehabilitation. 2012;19(6):463–70.

14. Saunders DH, Sanderson M, Brazzelli M, Greig CA, Mead GE. Physical fitness training for stroke patients. Cochrane Database Syst Rev. 2013(10):Cd003316.

15. Ivey FM, Hafer-Macko CE, Macko RF. Task-oriented treadmill exercise training in chronic hemiparetic stroke. J Rehabil Res Dev. 2008;45(2):249–59.

16. Ivey FM, Hafer-Macko CE, Ryan AS, Macko RF. Impaired leg vasodilatory function after stroke: adaptations with treadmill exercise training. Stroke. 2010;41(12):2913–7.

17. Ivey FM, Ryan AS, Hafer-Macko CE, Goldberg AP, Macko RF. Treadmill aerobic training improves glucose tolerance and indices of insulin sensitivity in disabled stroke survivors: a preliminary report. Stroke. 2007;38(10):2752–8.

18. Ivey FM, Ryan AS, Hafer-Macko CE, Macko RF. Improved cerebral vasomotor reactivity after exercise training in hemiparetic stroke survivors. Stroke. 2011;42(7):1994–2000.

19. Luft AR, Macko RF, Forrester LW, Villagra F, Ivey F, Sorkin JD, et al. Treadmill exercise activates subcortical neural networks and improves walking after stroke: a randomized controlled trial. Stroke. 2008;39(12):3341–50.

20. Macko RF, DeSouza CA, Tretter LD, Silver KH, Smith GV, Anderson PA, et al. Treadmill aerobic exercise training reduces the energy expenditure and cardiovascular demands of hemiparetic gait in chronic stroke patients. A preliminary report. Stroke. 1997;28(2):326–30.

21. Macko RF, Ivey FM, Forrester LW, Hanley D, Sorkin JD, Katzel LI, et al. Treadmill exercise rehabilitation improves ambulatory function and cardiovascular fitness in patients with chronic stroke: a randomized, controlled trial.Stroke. 2005;36(10):2206–11.

22. Macko RF, Smith GV, Dobrovolny CL, Sorkin JD, Goldberg AP, Silver KH. Treadmill training improves fitness reserve in chronic stroke patients. Archives of physical medicine and rehabilitation. 2001;82(7):879–84.

23. Jurkiewicz MT, Marzolini S, Oh P. Adherence to a home-based exercise program for individuals after stroke. Topics in stroke rehabilitation. 2011;18(3):277–84.

24. Moholdt T, Aamot IL, Granoien I, Gjerde L, Myklebust G, Walderhaug L, et al. Long-term follow-up after cardiac rehabilitation: a randomized study of usual care exercise training versus aerobic interval training after myocardial infarction. Int J Cardiol. 2011;152(3):388–90.

25. Tiedemann A, Sherrington C, Dean CM, Rissel C, Lord SR, Kirkham C, et al. Predictors of adherence to a structured exercise program and physical activity participation in community dwellers after stroke. Stroke Res Treat. 2012;2012:136525.

26. Duncan P, Studenski S, Richards L, Gollub S, Lai SM, Reker D, et al. Randomized clinical trial of therapeutic exercise in subacute stroke. Stroke. 2003;34(9):2173–80.

27. Kwakkel G. Impact of intensity of practice after stroke: issues for consideration. Disability and rehabilitation. 2006;28(13-14):823–30.

28. Boyne P, Dunning K, Carl D, Gerson M, Khoury J, Kissela B. High-intensity interval training in stroke rehabilitation. Topics in stroke rehabilitation. 2013;20(4):317–30.

29. Billinger SA, Boyne P, Coughenour E, Dunning K, Mattlage A. Does aerobic exercise and the FITT principle fit into stroke recovery? Curr Neurol Neurosci Rep. 2015;15(2):519.

30. Boyne P, Billinger S, MacKay-Lyons M, Barney B, Khoury J, Dunning K. Aerobic Exercise Prescription in Stroke Rehabilitation: A Web-Based Survey of US Physical Therapists. J Neurol Phys Ther. 2017;41(2):119–28.

31. Lau KW, Mak MK. Speed-dependent treadmill training is effective to improve gait and balance performance in patients with sub-acute stroke. J Rehabil Med. 2011;43(8):709–13.

32. Pohl M, Mehrholz J, Ritschel C, Rückriem S. Speed-dependent treadmill training in ambulatory hemiparetic stroke patients: a randomized controlled trial. Stroke. 2002;33(2):553–8.

33. Boyne P, Dunning K, Carl D, Gerson M, Khoury J, Rockwell B, et al. High-Intensity Interval Training and Moderate-Intensity Continuous Training in Ambulatory Chronic Stroke: Feasibility Study. Physical therapy. 2016;96(10):1533–44.

34. Boyne P, Scholl V, Doren S, Carl D, Billinger SA, Reisman DS, et al. Locomotor training intensity after stroke: Effects of interval type and mode. Topics in stroke rehabilitation. 2020:1–11.

35. Reisman D, Kesar T, Perumal R, Roos M, Rudolph K, Higginson J, et al. Time course of functional and biomechanical improvements during a gait training intervention in persons with chronic stroke. J Neurol Phys Ther. 2013;37(4):159–65.

36. Williams LS, Brizendine EJ, Plue L, Bakas T, Tu W, Hendrie H, et al. Performance of the PHQ-9 as a screening tool for depression after stroke. Stroke. 2005;36(3):635–8.

37. Fugl-Meyer AR, Jaasko L, Leyman I, Olsson S, Steglind S. The post-stroke hemiplegic patient. 1. a method for evaluation of physical performance. Scand J Rehabil Med. 1975;7(1):13–31.

38. Bohannon RW, Smith MB. Interrater reliability of a modified Ashworth scale of muscle spasticity. Physical therapy. 1987;67(2):206–7.

39. Brott T, Adams HP, Jr., Olinger CP, Marler JR, Barsan WG, Biller J, et al. Measurements of acute cerebral infarction: a clinical examination scale. Stroke. 1989;20(7):864–70.

40. Flansbjer UB, Holmback AM, Downham D, Patten C, Lexell J. Reliability of gait performance tests in men and women with hemiparesis after stroke. J Rehabil Med. 2005;37(2):75–82.

41. ATS statement: guidelines for the six-minute walk test. Am J Respir Crit Care Med. 2002;166(1):111–7.

42. Eng JJ, Dawson AS, Chu KS. Submaximal exercise in persons with stroke: test-retest reliability and concurrent validity with maximal oxygen consumption. Archives of physical medicine and rehabilitation. 2004;85(1):113–8.

43. Borg G. Perceived exertion as an indicator of somatic stress. Scand J Rehabil Med. 1970;2(2):92–8.

44. Pescatello LS, American College of Sports M. ACSM’s guidelines for exercise testing and prescription. Philadelphia: Wolters Kluwer/Lippincott Williams & Wilkins Health; 2014.

45. Boyne P, Reisman D, Brian M, Barney B, Franke A, Carl D, et al. Ventilatory threshold may be a more specific measure of aerobic capacity than peak oxygen consumption rate in persons with stroke. Topics in stroke rehabilitation. 2017;24(2):149–57.

46. Tucker CA, Escorpizo R, Cieza A, Lai JS, Stucki G, Ustun TB, et al. Mapping the content of the Patient-Reported Outcomes Measurement Information System (PROMIS(R)) using the International Classification of Functioning, Health and Disability. Qual Life Res. 2014;23(9):2431–8.

47. Mehrholz J, Wagner K, Rutte K, Meissner D, Pohl M. Predictive validity and responsiveness of the functional ambulation category in hemiparetic patients after stroke. Archives of physical medicine and rehabilitation. 2007;88(10):1314–9.

48. Golicki D, Niewada M, Buczek J, Karlinska A, Kobayashi A, Janssen MF, et al. Validity of EQ-5D-5L in stroke. Qual Life Res. 2015;24(4):845–50.

49. Salbach NM, Mayo NE, Hanley JA, Richards CL, Wood-Dauphinee S. Psychometric evaluation of the original and Canadian French version of the activities-specific balance confidence scale among people with stroke. Archives of physical medicine and rehabilitation. 2006;87(12):1597–604.

50. Jaeschke R, Singer J, Guyatt GH. Measurement of health status. Ascertaining the minimal clinically important difference. Control Clin Trials. 1989;10(4):407–15.

51. Ada L, Dean CM, Lindley R. Randomized trial of treadmill training to improve walking in community-dwelling people after stroke: the AMBULATE trial. Int J Stroke. 2013;8(6):436–44.

52. Dean CM, Ada L, Lindley RI. Treadmill training provides greater benefit to the subgroup of community-dwelling people after stroke who walk faster than 0.4m/s: a randomised trial. J Physiother. 2014;60(2):97–101.

53. Boyne P, Dunning K, Carl D, Gerson M, Khoury J, Kissela B. Within-session responses to high-intensity interval training in chronic stroke. Medicine and science in sports and exercise. 2015;47(3):476–84.

54. Boyne P, Meyrose C, Westover J, Whitesel D, Hatter K, Reisman DS, et al. Effects of Exercise Intensity on Acute Circulating Molecular Responses Poststroke. Neurorehabil Neural Repair. 2020;34(3):222–34.

55. Perera S, Mody SH, Woodman RC, Studenski SA. Meaningful change and responsiveness in common physical performance measures in older adults. J Am Geriatr Soc. 2006;54(5):743–9.

56. Guo Y, Logan HL, Glueck DH, Muller KE. Selecting a sample size for studies with repeated measures. BMC Med Res Methodol. 2013;13:100.

57. Fitzmaurice G, Laird N, Ware J. Applied longitudinal analysis. 2nd ed. Hoboken, NJ: John Wiley & Sons, Inc.; 2011.

58. Dobkin BH. Progressive Staging of Pilot Studies to Improve Phase III Trials for Motor Interventions. Neurorehabil Neural Repair. 2009;23(3):197–206.

59. Benjamini Y, Hochberg Y. Controlling the False Discovery Rate: A Practical and Powerful Approach to Multiple Testing. Journal of the Royal Statistical Society Series B (Methodological). 1995;57(1):289–300.

60. Dean CM, Rissel C, Sherrington C, Sharkey M, Cumming RG, Lord SR, et al. Exercise to enhance mobility and prevent falls after stroke: the community stroke club randomized trial. Neurorehabil Neural Repair. 2012;26(9):1046–57.

61. Hornby TG, Campbell DD, Kahn JH, Demott T, Moore JL, Roth HR. Enhanced gait-related improvements after therapist-versus robotic-assisted locomotor training in subjects with chronic stroke: a randomized controlled study. Stroke. 2008;39(6):1786–92.

62. Kim DK, Oh DW. Repeated Use of 6-min Walk Test with Immediate Knowledge of Results for Walking Capacity in Chronic Stroke: Clinical Trial of Fast versus Slow Walkers. Journal of stroke and cerebrovascular diseases : the official journal of National Stroke Association. 2019;28(11):104337.

63. Salbach NM, Mayo NE, Wood-Dauphinee S, Hanley JA, Richards CL, Côté R. A task-orientated intervention enhances walking distance and speed in the first year post stroke: a randomized controlled trial. Clin Rehabil. 2004;18(5):509–19.

64. Sullivan KJ, Knowlton BJ, Dobkin BH. Step training with body weight support: effect of treadmill speed and practice paradigms on poststroke locomotor recovery. Archives of physical medicine and rehabilitation. 2002;83(5):683–91.

65. Bowden MG, Behrman AL, Neptune RR, Gregory CM, Kautz SA. Locomotor rehabilitation of individuals with chronic stroke: difference between responders and nonresponders. Archives of physical medicine and rehabilitation. 2013;94(5):856–62.

66. Burke E, Dobkin BH, Noser EA, Enney LA, Cramer SC. Predictors and biomarkers of treatment gains in a clinical stroke trial targeting the lower extremity. Stroke. 2014;45(8):2379–84.

67. Dobkin BH, Nadeau SE, Behrman AL, Wu SS, Rose DK, Bowden M, et al. Prediction of responders for outcome measures of locomotor Experience Applied Post Stroke trial. J Rehabil Res Dev. 2014;51(1):39–50.

68. Freyssin C, Verkindt C, Prieur F, Benaich P, Maunier S, Blanc P. Cardiac rehabilitation in chronic heart failure: effect of an 8-week, high-intensity interval training versus continuous training. Archives of physical medicine and rehabilitation. 2012;93(8):1359–64.

69. Fu TC, Wang CH, Lin PS, Hsu CC, Cherng WJ, Huang SC, et al. Aerobic interval training improves oxygen uptake efficiency by enhancing cerebral and muscular hemodynamics in patients with heart failure. Int J Cardiol. 2013;167(1):41–50.

70. Guiraud T, Nigam A, Gremeaux V, Meyer P, Juneau M, Bosquet L. High-intensity interval training in cardiac rehabilitation. Sports Med. 2012;42(7):587–605.

71. Moholdt T, Aamot IL, Granoien I, Gjerde L, Myklebust G, Walderhaug L, et al. Aerobic interval training increases peak oxygen uptake more than usual care exercise training in myocardial infarction patients: a randomized controlled study. Clin Rehabil. 2012;26(1):33–44.

72. Moholdt TT, Amundsen BH, Rustad LA, Wahba A, Lovo KT, Gullikstad LR, et al. Aerobic interval training versus continuous moderate exercise after coronary artery bypass surgery: a randomized study of cardiovascular effects and quality of life. Am Heart J. 2009;158(6):1031–7.

73. Rognmo O, Hetland E, Helgerud J, Hoff J, Slordahl SA. High intensity aerobic interval exercise is superior to moderate intensity exercise for increasing aerobic capacity in patients with coronary artery disease. Eur J Cardiovasc Prev Rehabil. 2004;11(3):216–22.

74. Wisloff U, Stoylen A, Loennechen JP, Bruvold M, Rognmo O, Haram PM, et al. Superior cardiovascular effect of aerobic interval training versus moderate continuous training in heart failure patients: a randomized study. Circulation. 2007;115(24):3086–94.

75. Common terminology criteria for adverse events (CTCAE) version 4.0. National Cancer Institute. 2009.

76. Burgomaster KA, Hughes SC, Heigenhauser GJ, Bradwell SN, Gibala MJ. Six sessions of sprint interval training increases muscle oxidative potential and cycle endurance capacity in humans. J Appl Physiol (1985). 2005;98(6):1985–90.

77. Gibala MJ, Little JP, van Essen M, Wilkin GP, Burgomaster KA, Safdar A, et al. Short-term sprint interval versus traditional endurance training: similar initial adaptations in human skeletal muscle and exercise performance. J Physiol. 2006;575(Pt 3):901–11.

78. Hood MS, Little JP, Tarnopolsky MA, Myslik F, Gibala MJ. Low-volume interval training improves muscle oxidative capacity in sedentary adults. Medicine and science in sports and exercise. 2011;43(10):1849–56.

79. Bartlett JD, Hwa Joo C, Jeong TS, Louhelainen J, Cochran AJ, Gibala MJ, et al. Matched work high-intensity interval and continuous running induce similar increases in PGC-1alpha mRNA, AMPK, p38, and p53 phosphorylation in human skeletal muscle. J Appl Physiol (1985). 2012;112(7):1135–43.

80. Burgomaster KA, Howarth KR, Phillips SM, Rakobowchuk M, Macdonald MJ, McGee SL, et al. Similar metabolic adaptations during exercise after low volume sprint interval and traditional endurance training in humans. J Physiol. 2008;586(1):151–60.

81. Rakobowchuk M, Tanguay S, Burgomaster KA, Howarth KR, Gibala MJ, MacDonald MJ. Sprint interval and traditional endurance training induce similar improvements in peripheral arterial stiffness and flow-mediated dilation in healthy humans. Am J Physiol Regul Integr Comp Physiol. 2008;295(1):R236–42.

